# Assessment of a Super Acuity Test Chart for Hyperopia Screening

**DOI:** 10.64898/2026.04.24.26351668

**Authors:** Lene A. Hagen, Ellen Svarverud, Ieva Krastina, Graeme E. MacKenzie, Rigmor C. Baraas

**Author notes:** Corresponding authors: Rigmor C. Baraas, National Centre for Optics, Vision and Eye Care, Faculty of Health and Social Sciences, University of South-Eastern Norway, Hasbergs vei 36, 3616 Kongsberg, Norway., Graeme E. MacKenzie, Riemann Limited, Upper Heyford, Oxfordshire, United Kingdom……. Funding information: The study was funded by the European Union’s Horizon Europe research and innovation program under grant agreement No 101070155 (The Equitable, Inclusive, and Human-Centered XR Project [XR4Human]) and the University of South-Eastern Norway. Commercial relationships disclosures: L. A. Hagen (None); E. Svarverud (None); I. Krastina (None); G. E. MacKenzie (None); R. C. Baraas (None).

## Abstract

**Purpose:** To assess the repeatability of a prototype super acuity test chart for measuring visual acuity at 12.5 cm, and its ability to detect hyperopia in adolescents and young adults.

**Methods:** Repeatability was estimated as within-subject standard deviation of three repeated super acuity measurements performed in 41 university students (19–26 years). Associations between super acuity and cycloplegic refractive errors, ocular biometry, distance visual acuity, accommodation, age, and sex were assessed in 119 high school students (16–18 years) using linear mixed-effects models. ROC curves and Youden index were used to estimate the best super acuity thresholds to detect rest hyperopia.

**Results:** Mean super acuities in the university and high school cohorts were 0.14 ± 0.13 and 0.12 ± 0.11 logMAR, respectively. Repeatability was 0.031. Super acuity was poorer in those with uncorrected hyperopia [spherical equivalent refractive error (SER) ≥ 1.00 D] than the others [SER < 1.00 D; *P =* 0.039]. There were significant associations between poorer super acuity and more positive ametropia (SER; *P =* 0.026), poorer accommodation amplitude (*P <* 0.001), shorter axial length (*P =* 0.013), male sex (*P <* 0.001), and age (*P =* 0.037). Sensitivity and specificity for detecting hyperopia (SER ≥ 1.00 D) were 63.2% and 64.2%, respectively, at a super acuity threshold of 0.09 logMAR.

**Discussion:** The super acuity prototype shows promise as a screening indicator for hyperopia. Further studies are needed to optimize the test and testing protocol, and to assess its ability to detect uncorrected hyperopia in children.

## Introduction

Uncorrected refractive errors constitute a significant global public health concern.^1^ A well-documented challenge in vision screening, particularly in the detection of refractive errors, lies in the inadequacy of current tests to accurately identify hyperopia and distinguish it from myopia or emmetropia.^2^ Effective detection necessitates the use of cycloplegic agents to ensure the relaxation of accommodation.^3–5^ However, the application of cycloplegic drops is not only time-consuming but also requires the presence of authorized health care personnel, which is not always feasible and incurs additional costs. These constraints limit our ability to effectively identify individuals with uncorrected hyperopia.

Assessing visual acuity at 6 meters is a highly effective method for identifying individuals with myopia.^6^ Similarly, it would be advantageous to have a visual acuity chart for near vision that could help identify individuals with uncorrected or under-corrected hyperopia, a super acuity test chart. This study aimed to evaluate a prototype near visual acuity chart, designed in accordance with the principles of logMAR charts,^7^ for testing at 12.5 cm. The purpose of the short test distance is to introduce a high accommodation demand (8 dioptres (D) of accommodation is required for an emmetrope at 12.5 cm test distance). Uncorrected hyperopes will additionally need to accommodate to compensate for their refractive error, which leads to an even higher accommodation demand, depending on the magnitude of hyperopia. Greater accommodative stress may manifest as increased accommodative lag, or accommodative instability, that degrades the ability for optical resolution.^8, 9^ The mismatch between accommodative response and demand, and the resulting optical blur, may drive poorer super acuity performance of hyperopic eyes compared to their emmetropic or myopic counterparts.

The first prototype version of the super acuity test chart was created using Adobe InDesign and developed directly onto lithographic metal plates through a digital prepress method employed in offset printing to enhance precision. To maintain accuracy, laser-etched plates were utilized directly and housed within a 3D-printed frame specifically designed to ensure the testing distance of 12.5 cm.

When testing at such a close distance, several factors come into play, including differences in accommodation, spectacle magnification, and retinal image magnification associated with axial length.^10^ Additionally, eyes with longer axial lengths, typically associated with myopia, provide higher angular sampling density (increase in cones/deg²) in and around the fovea compared with emmetropic eyes, which generally have shorter axial lengths.^11^ This higher density offers the potential for better visual acuity.^12^ Moreover, the oculomotor system actively adjusts fixational drift behaviour, tending to move the stimulus towards retinal areas with higher cone densities. This oculomotor strategy, which maximizes the use of the most densely packed photoreceptors within the foveola, may also favour individuals with higher angular sampling density.^13^ Since angular cone density generally increases with axial length, shorter eyes may exhibit lower sampling density.^12^ Therefore, the higher accommodation demand and the shorter axial lengths, typically observed in hyperopic individuals, lead to the hypothesis that individuals who are uncorrected or under-corrected hyperopes may exhibit lower super acuity.

This study aimed to measure super acuity at 12.5 cm in adolescents and young adults with myopia, emmetropia, and hyperopia, along with associated measures of ocular biometry. The objectives were to describe the prototype version of the super acuity test, to assess the precision of the super acuity prototype in terms of within-session repeatability and between-visit reproducibility, and to evaluate the performance of the super acuity prototype for screening of rest hyperopia. Furthermore, the results are discussed in relation to possible improvements in the chart design and testing protocol, recognising that the present version is an initial prototype implementation and that further optimisation may improve sensitivity for detecting uncorrected hyperopia in younger populations, particularly children.

## Methods

### Participants

There were two cohorts in this study: (1) a university cohort, used for repeatability and reproducibility analyses; and (2) an upper secondary school (“high school”) cohort, used to examine associations with refractive error and to assess screening performance. All participants were recruited from the South-Eastern part of Norway. The study was approved by the Regional Committee for Medical and Health Research Ethics in South East Norway and followed the Declaration of Helsinki. Written consent was obtained after the participants had received oral and written information about the study, and prior to any measurements.

### Optometric Measurements

If the participant used glasses or contact lenses, they wore their habitual correction during all optometric measurements. Baseline optometric measurements were performed according to standard guidelines and included ocular preference, monocular distance logMAR visual acuity (VA) at 6 m, and monocular accommodation amplitude (mean of one push-in- and one pull-out-measurement with the Royal Air Force near-point rule).

Topical 1% cyclopentolate hydrochloride (Minims single use; Bausch & Lomb UK Ltd, Kingston upon Thames, UK) was used to obtain cycloplegic autorefraction (Huvitz HRK-8000A Auto-REF Keratometer; Huvitz Co. Ltd., Gyeonggi-do, Korea) and ocular biometry (Zeiss IOLMaster 700; Carl Zeiss Meditec AG, Jena, Germany). One drop of cyclopentolate was used for participants with blue and light green irides and two drops for those with dark green or brown irides. Cycloplegia was verified by the absence of the pupillary light reflex and accommodation amplitude of 2 D or less, typically observed 15–30 minutes after administering the drops. For autorefraction, the mean of five readings was used for further analyses. Ocular biometry included axial length (AL), corneal radius (CR), crystalline lens thickness (LT), and aqueous chamber depth (ACD). Measurements were performed by authorized optometrists and optometry students under supervision.

### The Super Acuity Test Chart

Super acuity was always measured before instillation of cycloplegic drops, with participants wearing their habitual correction. The super acuity test was conducted monocularly, with separate charts consisting of four rows of letter optotypes positioned in front of each eye. The test distance was 12.5 cm. To ensure the correct test distance, the chart was placed in a 3D-printed stand connected to a wearable spectacle frame suitable for those not wearing prescription glasses (Figure 1). For those wearing prescription glasses, the stand was held/secured at the side of the glasses in the spectacle plane to obtain the 12.5 cm test distance.

**Figure 1:**
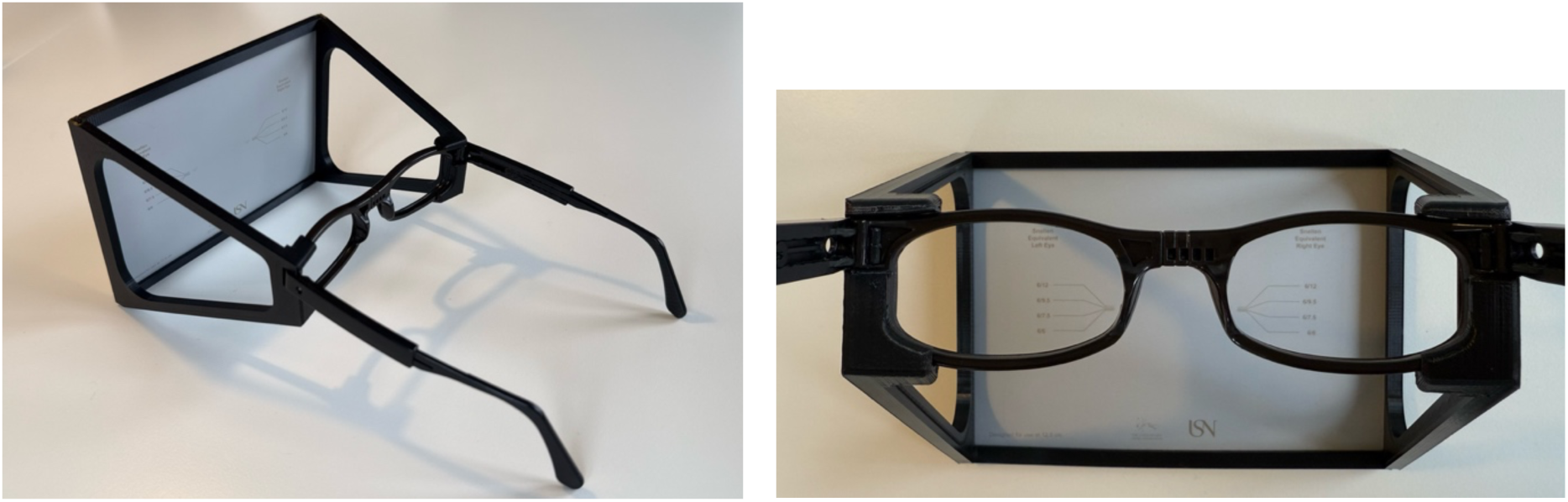
The super acuity test with the 3D-printed stand connected to a spectacle frame. The spectacle frame was removed for those wearing own prescription glasses.

The charts were logMAR charts, with five letter optotypes on each letter row and with each successive row decreasing in size by a constant size progression ratio of 10^0.1^ (≈ 1.259).^7^ The distance between adjacent letters was equal to the width of the letters in the respective row, and the spaces between successive rows corresponded to the height of the letters in the smaller row.^7^ This corresponded to a logMAR visual acuity (VA) of 0.30, 0.20, 0.10 and 0.00 for the four rows of letter optotypes, with each letter corresponding to a value of 0.02 logMAR units. Eleven different sans-serif 5 × 5 height-to-width ratio letter optotypes were presented in the two test charts. An overview of the optotype letters in each test chart is given in Table 1.

**Table 1.**
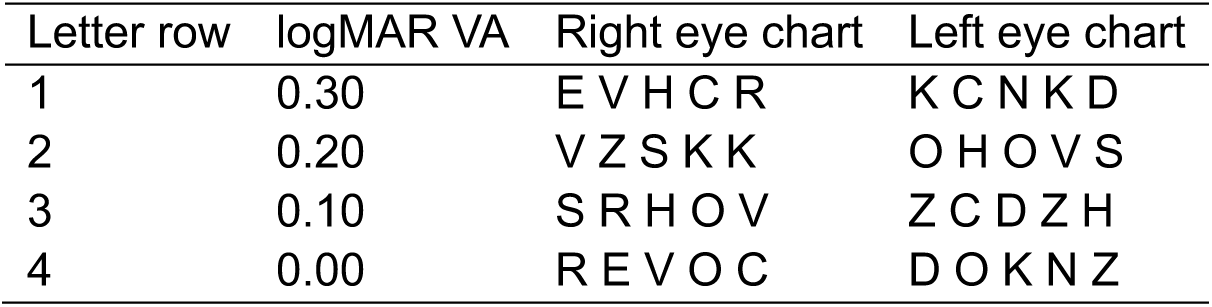
An overview of optotype letters presented in each letter row of the super acuity test charts.

To create the test chart, a vector graphic of the chart was created in Adobe InDesign, allowing design precision down to 1 micron. The charts were developed directly onto lithographic metal plates using the Computer-to-Plate (CtP) process. The vector graphic file was loaded into a CtP system, where a laser etched the chart design onto a photosensitive metal plate. Only the optotype areas were exposed, making these sections ‘ink-receptive’ and forming the exact chart shapes on the plate itself. After etching, the plate went through a development step to remove any unexposed coating, leaving only the optotype shapes. Ordinarily, these plates would be inked to produce thousands of paper copies via offset printing, but inking was bypassed to avoid potential inaccuracies. Instead, the laser-etched plates were used directly for greater precision. At the time of production, the etching process could in principle produce optotypes smaller than logMAR 0.00 at 12.5 cm, but not with sufficient consistency. When attempting to manufacture logMAR 0.00 optotypes for a 10 cm test distance, approximately half of the letters showed inconsistencies in limb spacing. In practice, etching gaps or lines narrower than 0.036 mm (the bar/space width for a logMAR 0.00 optotype at 12.5 cm) led to an unacceptably high defect rate, and therefore the final chart was limited to logMAR 0.00 at 12.5 cm.

### Procedure for Measuring Super Acuity

The participant wore their habitual correction during the test. The test was performed monocularly with the contralateral eye covered, starting with the right eye. The participant was instructed to read each letter out loud starting at the left of the top row and encouraged to proceed to their recognition threshold, including guesses for the letters around their threshold. Each letter read was recorded, and those not identified were marked as not seen. If the participant changed their mind about an optotype, the final response was recorded. Errors made above the line of threshold acuity was included in the calculation of the super acuity result. The chart illumination was in the range of 750–1250 lux.

### Reproducibility and Repeatability of Super Acuity Measurements

Measurements of reproducibility and repeatability of the super acuity test were conducted on a separate day, in which the sample of university students participated. The criterion for inclusion of follow-up measurements was willingness to participate a second time.

To reproduce the test situation as similar as possible to the baseline super acuity measurements, a set of optometric measurements were conducted prior to measuring super acuity, including monocular distance logMAR VA, monocular accommodation amplitude, and stereoacuity at 40 cm. Super acuity measurements were conducted under identical conditions three times for each eye, and by alternating measurements of the right and left eye, starting with the right eye.

Between-visit reproducibility of super acuity was evaluated by comparing the baseline measurement on the first visit with the first follow-up measurement conducted on the second visit. Within-session repeatability of super acuity was evaluated using the three consecutive measurements performed under identical conditions on the second visit.

### Validation of the Optotype Size

To validate the size of the optotypes, the optotypes were imaged by a Hitachi SU3500 Scanning Electron Microscope (FlexSEM 1000; Hitachi High-Technologies Corporation, Tokyo, Japan) at a magnification of 170–371× (Figure 2). The full vertical and horizontal size of the optotypes [in microns] were measured from measurement lines that were aligned by a trained microscopist and used to estimate the logarithm of the minimum angle of resolution (logMAR) of each optotype.

**Figure 2.**
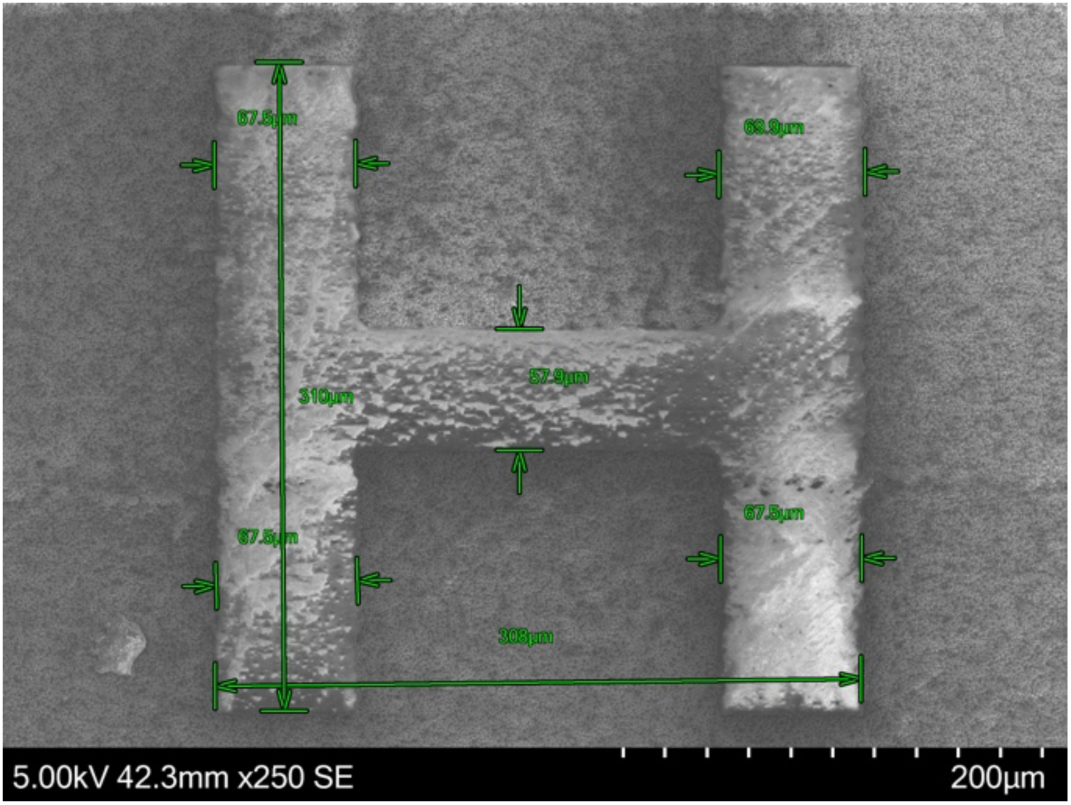
Image from the Hitachi SU3500 Scanning Electron Microscope of an optotype letter (H) with measurement lines aligned by a trained microscopist. This optotype letter is from row 2 of the left eye super acuity test chart, assumed to represent logMAR 0.20. Expected full height and width for an optotype representing logMAR 0.20 at 12.5 cm test distance is 288.1 microns (= 0.2881 mm).

Two optotypes per row, one from the left and one from the right eye chart, on two randomly chosen super acuity test plates, were validated. Table 2 provides an overview of mean logMAR from the measured optotype size compared with the expected size for each letter row. The mean difference between measured and expected logMAR ranged from 0.003 to 0.012 logMAR. Measured letter heights deviated from the intended logMAR values by at most ∼0.01 log units, which is within the ±0.01 logMAR (≈2%) manufacturing tolerance recommended by the ICO Visual Acuity Measurement Standard.^14^ Ratio of size progression from one optotype row to the next ranged from 1.239 to 1.277, which is within ±2% of the expected size progression ratio in a logarithmic chart (10^0.1^ ≈ 1.259).^7^

**Table 2.** Validation of optotype letter size.

| Letter row | Expected logMAR VA | Number of letters controlled<br>n | Measured logMAR<br>Mean ± SD | Difference in logMAR<br>Mean ± SD | Size progression ratio |
| --- | --- | --- | --- | --- | --- |
| 1 | 0.30 | 4 | 0.309 ± 0.005 | 0.009 ± 0.005 | 1.277 |
| 2 | 0.20 | 4 | 0.203 ± 0.003 | 0.003 ± 0.003 | 1.252 |
| 3 | 0.10 | 4 | 0.105 ± 0.006 | 0.005 ± 0.006 | 1.239 |
| 4 | 0.00 | 3* | 0.012 ± 0.003 | 0.012 ± 0.003 | Last line of the chart |

\* For letter row 4, measurements of only 3 letter optotypes were available

### Data Analyses

Cycloplegic autorefraction was used to categorize ametropia as myopia (SER ≤ -0.50 D), hyperopia (SER ≥ 1.00 D), and astigmatism (absolute cylinder ≥ 0.75 D). The frequency of astigmatism was estimated in two ways: (1) for the whole sample (astigmatism overall), and (2) for those with no other ametropia, defined as SER in the region of emmetropia (-0.50 D < SER < 1.00 D; astigmatism only). Anisometropia was defined as a difference in SER ≥ 1.00 D between right and left eye. Rest ametropia was defined as the difference between the habitual prescription and each participant’s cycloplegic autorefraction result, since habitual correction was worn during measurements. These calculations were performed using power vectors.^15^

The statistical analyses were performed using R version 4.5.2,^16^ and statistical significance was defined as *P <* 0.05. Precision of the super acuity test was assessed by within-session repeatability (*S*_*r*_) for three repeated measurements performed without breaks, and by the repeatability limit (*r*), defined as 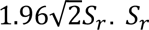 was estimated as the within-subject standard deviation derived from a one-way analysis of variance (ANOVA). The between-visit reproducibility analysis assessed reproducibility (*S_R_*) for the baseline measurement compared with the first measurement on the second visit, together with the corresponding reproducibility limit (*R*). Agreement between these measurements was assessed using Bland-Altman analysis, reported as the mean difference (bias) and the 95% limits of agreement (mean difference ± 1.96 × standard deviation of the difference).^17^ Repeatability was assessed in 41 university students. Using the McAlinden ANOVA approach, with 41 participants and three repeated measurements (*df* = 82), the relative standard error of the within-subject variance, and hence of the repeatability estimate, was approximately 15%, which was considered acceptable for a development study.^18^ Since baseline measurements with the same prescription correction were not available for eight of the university students, between-visit reproducibility was assessed in 33 participants. Using the McAlinden ANOVA approach, with 33 participants and two visits (*df* = 33), the relative standard error of the between-visit variance, and hence of the reproducibility estimate, was approximately 24%, which was considered acceptable for a development study, albeit less precise than the repeatability estimate.

Distributions of the data were checked using histograms and Q-Q plots. Kruskal-Wallis and non-parametric Wilcoxon rank-sum test were used to compare super acuity results across rest ametropia groups, since the assumption of normality was not met.

A linear mixed-effect model was used to examine the association between super acuity as the dependent variable and rest ametropia, ocular biometry, distance visual acuity, accommodation amplitude, age, and sex as independent variables. The model was fitted using the lmerTest package,^19^ which applies Satterthwaite’s correction to estimate *P*-values and degrees of freedom for the fixed effects. Confidence intervals were calculated using the Wald method. Measurements from both eyes were included in the analysis, and to account for this, the model included a random intercept for participant. Collinearity among the fixed-effect predictors was examined using variance inflation factors (VIFs); all retained predictors had VIF values below 3.4, suggesting acceptable collinearity. Two predictors, AL and CR, showed moderate collinearity (VIF = 3.34 and 3.14); of these, only AL was retained in the final model on the basis of better model fit. Sensitivity analyses were performed with SER and AL entered separately, and these yielded results comparable with those of the final model. Model assumptions were assessed by inspection of residuals and Q–Q plots. Because super acuity was bounded and included a small number of floor (0.00) and ceiling (0.40) logMAR values, a sensitivity analysis excluding these cases was also performed and yielded results comparable with those of the final model.

Receiver operating characteristic (ROC) curves and Youden’s Index^20^ were used to estimate optimal thresholds for screening of rest hyperopia, and sensitivity and specificity were calculated for these thresholds. The ROC analyses were conducted using one eye per participant, selected as the eye with the most hyperopic meridional power.

## Results

### Repeatability and Reproducibility (University Cohort)

For the study of repeatability, 41 healthy university students (83% females) aged 19–26 years (21.3 ± 1.9 yrs) participated. Table 3 provides an overview of the university cohort. The measurements of super acuity were performed with habitual prescription correction, and on the test day, 54% wore glasses and 7% wore contact lenses. Twenty-eight (68%) of dominant eyes and 23 (56%) of non-dominant eyes were emmetropic or fully corrected (i.e., had no rest ametropia).

**Table 3.**
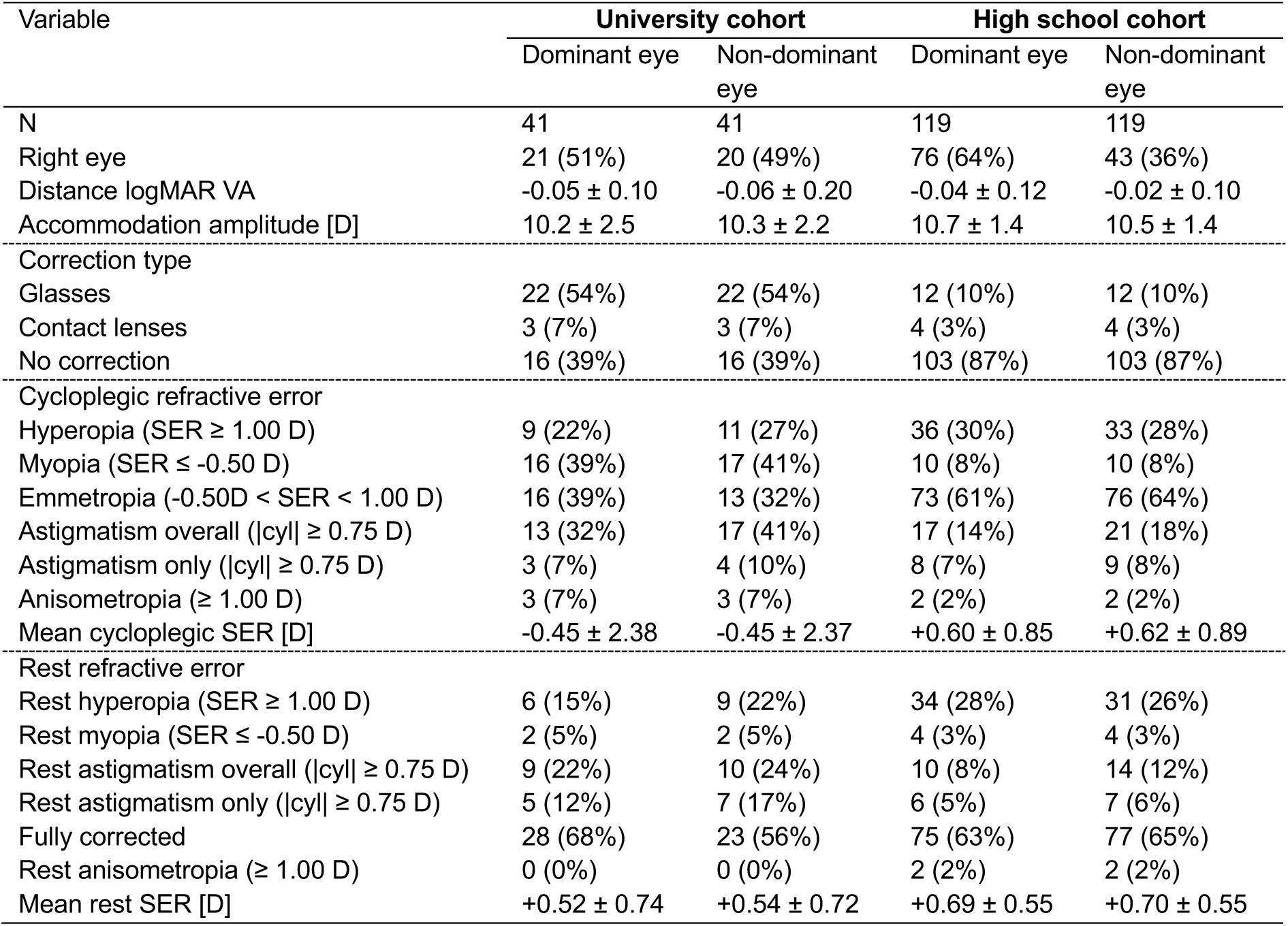
Overview of the university cohort [n = 41 (83% females); aged 19–26 years (21.3 ± 1.9 yrs)] and the high school cohort [n = 119 (45% females); aged 16–18 years (16.8 ± 0.8 yrs)].

Super acuity results for the three repeated measurements of the university cohort are presented in Table 4 and Figure 3. The mean ± SD of the three repeated measurements was 0.14 ± 0.13 logMAR in the dominant eye and 0.13 ± 0.12 logMAR in the non-dominant eye. Repeatability values [*S*_*r*_(*r*)] were 0.031 (0.085) and 0.039 (0.109) for the dominant and non-dominant eye, respectively. No correlations were found, in either eye, between each individual’s repeatability (*S*_*r*_) and super acuity, accommodation amplitude, cycloplegic refractive error, ametropia, or age (all *R²* < 0.05; all *P >* 0.05). Exclusion of the participants with super acuity values of 0.00 (all letters correctly read; n = 9) or 0.40 (no letters correctly read; n = 5), in one or more of the repeated measurements in the dominant eye, did not significantly change the repeatability value (*S_r_* = 0.035 for n = 27 vs. *S_r_* = 0.031 for n = 41).

**Table 4.** Overview of super acuity for the three repeated measurements (Rep 1, 2, and 3) for the university cohort (n = 41). Repeatability value (*S_r_*), and repeatability limit (*r*) were evaluated using the three consecutive measurements on the second visit.

| Eye | Three repeated measurements of super acuity | | | | Repeatability $S_r$ ($r$) |
| --- | --- | --- | --- | --- | --- |
| | Rep 1<br>Mean ± SD | Rep 2<br>Mean ± SD | Rep 3<br>Mean ± SD | Mean of all 3<br>Mean ± SD | |
| Dominant | 0.144 ± 0.135 | 0.132 ± 0.132 | 0.130 ± 0.132 | 0.135 ± 0.131 | 0.031 (0.085) |
| Non-dominant | 0.145 ± 0.123 | 0.129 ± 0.125 | 0.121 ± 0.121 | 0.132 ± 0.119 | 0.039 (0.109) |
| Right | 0.152 ± 0.125 | 0.140 ± 0.123 | 0.141 ± 0.122 | 0.144 ± 0.120 | 0.033 (0.092) |
| Left | 0.137 ± 0.133 | 0.122 ± 0.133 | 0.111 ± 0.130 | 0.123 ± 0.129 | 0.037 (0.103) |

**Figure 3.**
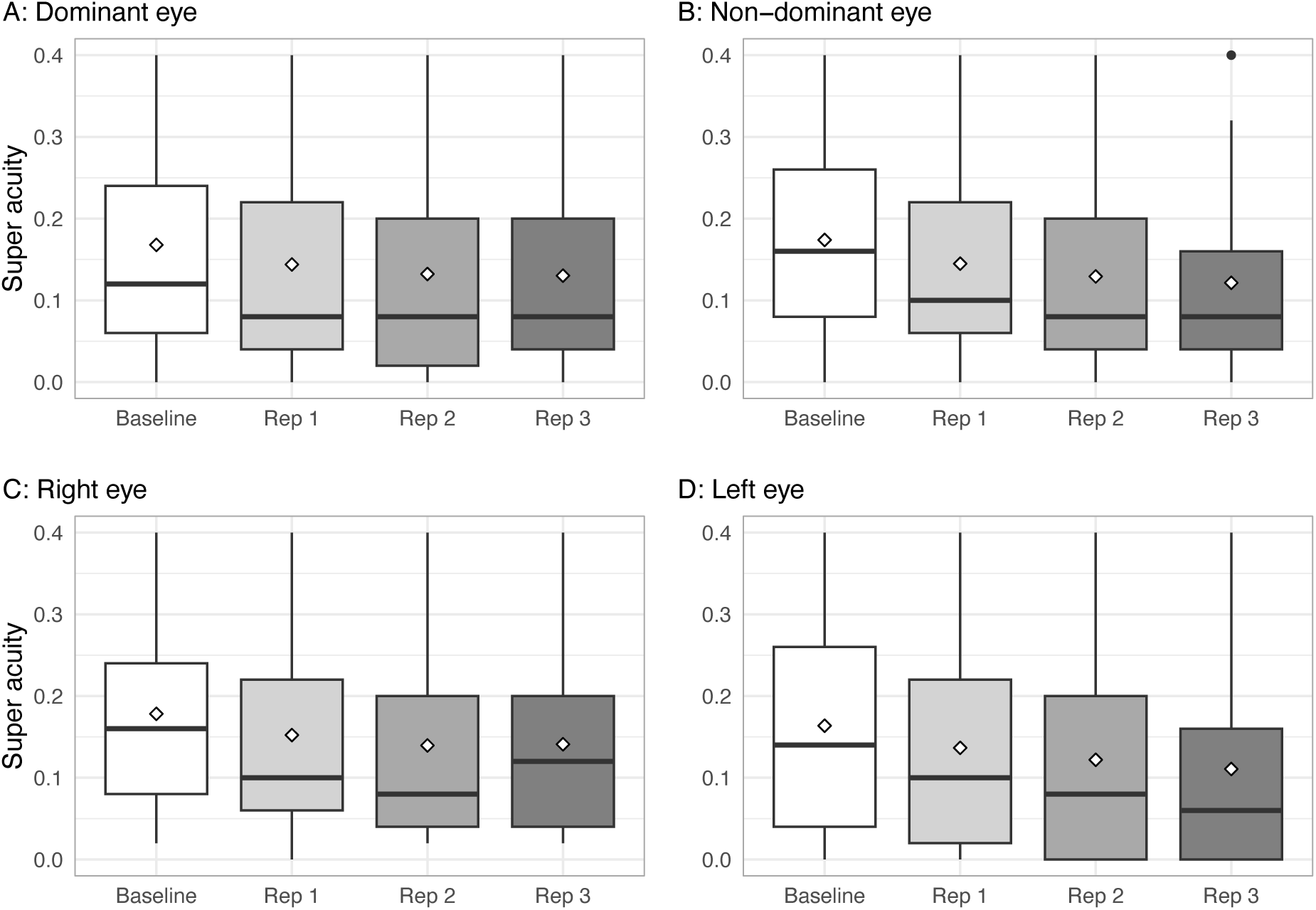
Boxplots showing super acuity for baseline (n = 33) and each of the three repeated measurements (Rep 1, 2, and 3; n = 41) in the (A) dominant, (B) non-dominant, (C) right, and (D) left eye of the university cohort. Diamonds and horizontal lines represent mean and median values, respectively.

A Bland-Altman analysis was used to assess the agreement between baseline super acuity and the first follow-up measurement on the second visit for all participants in the university cohort who used the same prescription correction at first (baseline) and second visit [n = 33 (79% females), aged 19–26 years (21.4 ± 2.0 yrs)]. Super acuity levels are presented in Table 5, and Bland-Altman plots are presented in Figure 4. Mean differences (95% limits of agreement) for the dominant and non-dominant eye were -0.04 ± 0.09 (-0.22–0.15) and -0.04 ± 0.08 (-0.19–0.12), respectively, indicating a slight improvement in super acuity from baseline to the first follow-up measurement. The between-visit reproducibility value (*S_R_*; within-subject standard deviation) and limit (*R*) for the dominant eye were 0.070 and 0.195, respectively.

**Table 5.** Overview of baseline and first follow-up measurement of super acuity for all participants in the university cohort who used the same prescription correction at first (baseline) and second visit (n = 33). Reproducibility value (*S_R_*) and reproducibility limit (*R*) were evaluated by comparing the baseline measurement with the first follow-up measurement conducted on the second visit.

| Eye | Super acuity | | Reproducibility<br>$S_R$ ($R$) |
| --- | --- | --- | --- |
| | Baseline<br>Mean ± SD | First follow-up<br>Mean ± SD | |
| Dominant | 0.168 ± 0.122 | 0.130 ± 0.127 | 0.070 (0.195) |
| Non-dominant | 0.174 ± 0.125 | 0.136 ± 0.121 | 0.061 (0.169) |

**Figure 4.**
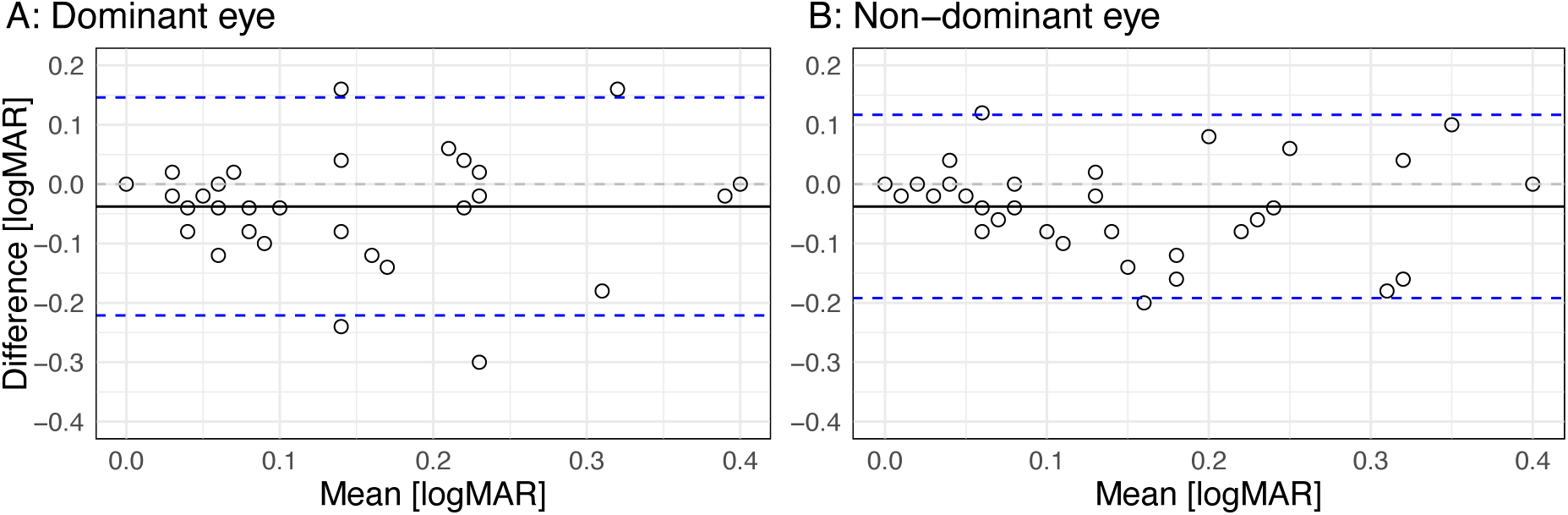
Bland-Altman plots comparing baseline super acuity with the first follow-up measurement conducted on the second visit for (A) the dominant and (B) the non-dominant eye of participants in the university cohort who used the same prescription correction at first and second visit (n = 33). The plots show the mean of the two super acuity measurements on the x-axis and their difference on the y-axis. The solid line represents the mean difference (bias), and the dashed lines indicate the 95% limits of agreement (mean ± 1.96 × SD).

In Table 6, mean distance VA and repeated super acuity in the dominant eye are summarized in categories of ametropia and rest ametropia. A comparison of super acuity across the four groups of rest ametropia, did not reveal a significant difference among the groups [Kruskal-Wallis; *χ²*(3) = 7.60, *P =* 0.055]. When comparing those with rest hyperopia (rest SER ≥ 1.00 D; n = 6) and the others (rest SER < 1.00 D; n = 35), those with rest hyperopia had poorer repeated super acuity than the others [*W* = 37, *P =* 0.013].

**Table 6.** Overview of distance visual acuity (VA) and super acuity in the dominant eye, categorized by ametropia and rest ametropia, for the university cohort (n = 41) and the high school cohort (n = 119).

|  | The university cohort |  |  | The high school cohort |  |  |
| --- | --- | --- | --- | --- | --- | --- |
|  | n | Distance VA | Repeated super acuity | n | Distance VA | Baseline super acuity |
| | | Mean ± SD | Mean ± SD | | Mean ± SD | Mean ± SD |
| All | 41 | -0.05 ± 0.10 | 0.14 ± 0.13 | 119 | -0.04 ± 0.12 | 0.12 ± 0.11 |
| Hyperopia | 9 | -0.10 ± 0.07 | 0.28 ± 0.12 | 36 | -0.06 ± 0.08 | 0.16 ± 0.13 |
| Myopia | 16 | -0.01 ± 0.10 | 0.11 ± 0.10 | 10 | 0.13 ± 0.30 | 0.07 ± 0.07 |
| Emmetropia | 16 | -0.06 ± 0.11 | 0.08 ± 0.11 | 73 | -0.05 ± 0.07 | 0.10 ± 0.09 |
| Rest hyperopia | 6 | -0.09 ± 0.05 | 0.26 ± 0.13 | 34 | -0.06 ± 0.07 | 0.16 ± 0.13 |
| Rest myopia | 2 | 0.17 ± 0.04 | 0.05 ± 0.04 | 4 | 0.34 ± 0.40 | 0.08 ± 0.06 |
| Rest astigmatism only | 5 | 0.02 ± 0.08 | 0.19 ± 0.19 | 6 | 0.04 ± 0.05 | 0.08 ± 0.04 |
| Fully corrected | 28 | -0.07 ± 0.10 | 0.10 ± 0.10 | 75 | -0.06 ± 0.07 | 0.10 ± 0.09 |

### The Association between Super Acuity and Rest Ametropia (High School Cohort)

The association between super acuity and rest ametropia was further assessed in a sample of upper secondary school students (high school cohort). Of the 121 upper secondary school students who participated, two were excluded from the analysis: one due to missing data and one due to high, uncorrected astigmatism in one eye (rest-cylinder = -3.87 D). The analysis included 119 adolescents (45% females) aged 16–18 years (16.8 ± 0.8 yrs). Table 3 provides an overview of the high school cohort. On the test day, 10% wore glasses and 3% wore contact lenses. Seventy-five (63%) of dominant eyes and 77 (65%) of non-dominant eyes were emmetropes or fully corrected.

An overview of distance visual acuity and super acuity for the dominant eye of the high school cohort is given in Table 6. Mean distance visual acuity was -0.04 ± 0.12 logMAR, and mean super acuity was 0.12 ± 0.11 logMAR. In the dominant eye, rest hyperopia was present in 29%, rest myopia was present in 3%, and rest astigmatism only (i.e., no other rest-ametropia) was present in 5%. As for the university cohort, those who had rest hyperopia in the dominant eye in the high school cohort (rest SER ≥ 1.00 D; n = 34) had poorer baseline super acuity (*W* = 1095.5, *P =* 0.039) than the other (rest SER < 1.00 D; n = 85).

A linear mixed-effects model with super acuity as the dependent variable was fitted to the full high school cohort dataset, including data from both eyes for each participant. A random intercept for the participant ID was added to account for the inclusion of both eyes. Details are presented in Table 7. The model showed significant associations between super acuity and rest ametropia (*β* = 0.030, *P =* 0.026), accommodation amplitude (*β* = -0.019, *P <* 0.001), axial length (*β* = -0.029, *P =* 0.013), sex (*β* = -0.062, *P <* 0.001), and age (*β* = 0.021, *P =* 0.037). This indicated poorer super acuity with a more positive rest ametropia, poorer accommodation amplitude, shorter axial length, and higher age, as well as poorer super acuity for males versus females. Due to the moderate correlation between axial length and rest ametropia (Pearson’s *r* = -0.34, 95% CI [-0.44, -0.22], *P* < 0.001), sensitivity analyses of the model were performed with axial length and rest ametropia entered separately. Although effect estimates varied slightly from the final model, the overall pattern and interpretation of results remained unchanged. Sensitivity analyses excluding boundary values of super acuity, that is, 16 eyes with super acuity = 0.00 and 10 eyes with super acuity = 0.40, yielded effect estimates of similar direction and magnitude to those of the final model, although two predictors, ametropia and age, no longer reached statistical significance. An interaction between axial length and sex was also tested but not found to be significant (*β* = 0.018, standard error (SE) = 0.022, *t* = 0.824, *P =* 0.412).

**Table 7.** Linear mixed effect model regression estimates for predictors of super acuity.

| Fixed effects | Estimate<br>($\beta$) | Standard<br>Error (SE) | 95% CI | $t$ | $P$-value |
| --- | --- | --- | --- | --- | --- |
| Intercept | 0.654 | 0.314 | 0.038 – 1.269 | 2.082 | 0.039 |
| Rest ametropia (SER) | 0.030 | 0.013 | 0.004 – 0.055 | 2.244 | 0.026 |
| Accommodation amplitude | -0.019 | 0.004 | -0.028 – (-0.011) | -4.405 | < 0.001 |
| Sex (female)* | -0.062 | 0.017 | -0.096 – (-0.028) | -3.598 | < 0.001 |
| Axial length | -0.029 | 0.012 | -0.051 – (-0.006) | -2.505 | 0.013 |
| Age | 0.021 | 0.010 | 0.002 – 0.040 | 2.112 | 0.037 |
| Random effects | Variance | Standard<br>deviation |  |  |  |
| Participant ID (intercept) | 0.007 | 0.083 |  |  |  |
| Model fit | Marginal | Conditional |  |  |  |
| $R$-squared | 0.23 | 0.83 | | | |

\* Reference category: male
Confidence intervals (CIs) have been calculated using the Wald method, and $P$ -values for fixed effects were calculated using Satterthwaite's approximations.

### Rest Hyperopia Screening Performance (High School Cohort)

For each individual in the high school cohort, data from the eye with the most hyperopic meridional power were used to assess the performance of the super acuity test for screening of rest hyperopia. First, the variable “VA-difference” was defined as the difference between distance visual acuity and super acuity (VA-difference = distance VA – super acuity). Then, ROC curves and Youden’s Index (Youden, 1950) were used to estimate the optimal threshold of (i) super acuity, (ii) VA-difference, and (iii) accommodation amplitude for screening of rest hyperopia at two levels (SER ≥ 1.00 D and SER ≥ 1.50 D). Results are presented in Table 8. For detecting rest hyperopia defined as SER ≥ 1.00 D (n = 38 of 119), the sensitivity and specificity were 63.2% and 64.2%, respectively, at a super acuity threshold of 0.09 logMAR or higher, and 44.7% and 74.1%, respectively, at a VA-difference of -0.19 logMAR or more negative. The optimal threshold for accommodation amplitude was 9.6 D or poorer, which gave a sensitivity and specificity of 47.4% and 77.8%. For detecting rest hyperopia defined as SER ≥ 1.50 D (n = 9 of 119), the sensitivity and specificity were 100.0% and 60.0%, respectively, at a super acuity threshold of 0.09 logMAR or higher.

**Table 8.** Estimated threshold values of (i) super acuity, (ii) VA-difference (difference between distance visual acuity and super acuity), and (iii) monocular accommodation amplitude, along with the sensitivity and specificity of detecting rest hyperopia at two levels (rest SER ≥ 1.00 D or rest SER ≥ 1.50 D).

|  | Threshold | Sensitivity [%] | Specificity [%] |
| --- | --- | --- | --- |
| **Rest hyperopia (SER ≥ 1.00 D, n = 38 of 119)** |  |  |  |
| Super acuity [logMAR] | $\geq 0.09$ | 63.2 | 64.2 |
| Difference in VA [logMAR] | $\leq -0.19$ | 44.7 | 74.1 |
| Monocular accommodation amplitude [D] | $\leq 9.6$ | 47.4 | 77.8 |
| **Rest hyperopia (SER ≥ 1.50 D, n = 9 of 119)** |  |  |  |
| Super acuity [logMAR] | $\geq 0.09$ | 100.0 | 60.0 |
| Difference in VA [logMAR] | $\leq -0.19$ | 88.9 | 72.3 |
| Monocular accommodation amplitude [D] | $\leq 11.1$ | 88.9 | 37.3 |

## Discussion

In this study, a prototype super acuity test for measuring near visual acuity at 12.5 cm was evaluated as a potential screening indicator for rest hyperopia. Precision of the super acuity test was assessed in terms of within-session repeatability and between-visit reproducibility in a group of university students, while the test’s performance for screening of rest hyperopia was evaluated in a group of upper secondary school students. The prototype demonstrated good repeatability and encouraging, but still moderate, screening performance for rest hyperopia in adolescents and young adults.

The within-session repeatability results of 0.085 (dominant eye) and 0.109 (non-dominant eye) for the super acuity prototype (Table 4) correspond to about 4 and 5.5 letters, respectively. This is in the same range as previously reported for distance visual acuity charts, such as about 4 letters for the Early-Treatment Diabetic Retinopathy Study (ETDRS) chart in the study of Veselý and Synek^21^, about 6–7 letters for the ETDRS chart in the study of Laidlaw, et al.^22^, about 4.5 letters for the Bailey-Lovie chart in the study of Ravikumar, et al.^23^, and about 7–8 letters for the ETDRS chart measured one month apart in a group of 6–11 year old myopic children in the study of Manny, et al.^24^. For computerised logMAR visual acuity measurement systems such as COMPlog, within-session repeatability has been reported to be approximately 5 letters^22^ and between-visit reproducibility about 6 letters^25^. The between-visit reproducibility of 0.195 in the dominant eye for the super acuity prototype in the current study (Table 5) corresponds to about 9–10 letters, which is within the upper end of what is reported for distance visual acuity charts and newer digital tools. Nevertheless, these early results are promising, given that the testing distance of 12.5 cm for the super acuity test is more demanding in terms of accommodation (8 D for an emmetrope) than traditional visual acuity testing performed at distance (around 6 m; a minimum of accommodation required for an emmetrope) or near (around 40 cm; 2.5 D accommodation required for an emmetrope). The contrast of the optotypes against the background of the super acuity test (Figure 1; laser-etched optotypes on a grey metal plate) is also lower than for standard white-background charts.

The super acuity prototype was designed to stress the accommodative system in individuals with uncorrected or under-corrected hyperopia and is expected to induce optical blur when there is a mismatch between the accommodative demand and response. Indeed, the super acuity for those with rest hyperopia was poorer than for the others both in the university and the high school cohorts (Table 6). Poorer super acuity was associated with a more positive rest ametropia (i.e., uncorrected hyperopia under habitual correction), poorer accommodation amplitude, shorter axial length, higher age, and male sex in the high school cohort (Table 7). Shorter eyes are associated with hyperopia and may exhibit lower angular sampling density in and around the fovea^12^, which could contribute to poorer super acuity, although the present data do not allow the underlying mechanism to be determined. The association between higher age and poorer super acuity may reflect age-related differences in accommodation^26^, but the age range in the high school cohort was narrow (16 to 18 years), and this finding should therefore be interpreted cautiously and confirmed in a broader sample. Super acuity was also found to be slightly better in females than males. The underlying reasons for this are likely multifactorial, and the clinical relevance is uncertain.

The super acuity test showed moderate screening performance for rest hyperopia in adolescents and young adults (Table 8), suggesting that the super acuity prototype may have value as a screening indicator for rest hyperopia. The sensitivity for super acuity (63.2%) for detecting rest hyperopia of ≥ 1.00 D was slightly better than for monocular accommodation amplitude (47.4%) and for VA-difference (44.7%), although the specificity was slightly poorer. Screening performance was therefore only moderate at the ≥ 1.00 D threshold. However, performance appeared stronger at the higher threshold of SER ≥ 1.50 D, particularly in terms of sensitivity, although this estimate was based on a very small number of participants and should therefore be interpreted cautiously. Even so, this raises the possibility that the prototype may be more effective at identifying higher levels of rest hyperopia, which may be more clinically or functionally relevant. More broadly, these findings should be interpreted in the context of an unresolved issue in paediatric vision care: the threshold at which hyperopia becomes functionally significant remains uncertain, particularly for outcomes such as reading, sustained near work, and academic performance. This uncertainty is further compounded by the practical difficulty of using cycloplegia in large-scale screening programmes.

Since the super acuity results reflect individual variations in accommodation ability, it is reasonable to assume that some individuals with rest hyperopia under habitual correction may still demonstrate good super acuity if their accommodation ability is high. Incorporating a time-related element in the super acuity testing paradigm, such as sustained viewing, repeated cycles, or limited time to failure, may improve the sensitivity and specificity for hyperopia screening. More broadly, the present findings suggest that both the chart design and the testing protocol remain open to optimisation, and that the current version should be regarded as a first prototype implementation rather than an optimised final design. To detect all individuals with uncorrected hyperopia, the use of cycloplegic agents may still be necessary to achieve full control of the accommodation. Nevertheless, the findings here suggest that near acuity measured with the super acuity prototype may have value as a screening indicator for rest hyperopia.

To our knowledge, this is the first near visual acuity test chart in which the resolution of the optotype letters gives the possibility to test at such a close distance (12.5 cm), introducing a high accommodative demand (8 D for an emmetrope). The presentation of the test chart in a stand connected to a spectacle frame ensured the correct test distance. Repeatability and reproducibility were explicitly evaluated using repeated measurements under similar conditions both on the same visit as well as with re-testing another day, allowing for quantification of both within-subject and between-session variability. It is a strength that the screening performance of the super acuity prototype for rest hyperopia was tested in a large sample of upper secondary school students, in which both cycloplegic autorefraction and ocular biometry were measured, even though the low frequency of hyperopia and astigmatism in the sample was a limitation.

Further research is needed to assess the screening performance of the super acuity prototype in children and younger adolescents, in larger samples with a higher frequency of uncorrected hyperopia, and to refine the test design and testing protocol with the aim of improving its performance as a screening indicator.

As hyperopia, if left uncorrected, may lead to amblyopia and strabismus^27^ and reduced academic performance,^28^ it is important to develop effective approaches to hyperopia screening. This study provides encouraging early evidence that the super acuity prototype may have potential as a practical and easy-to-use screening indicator for rest hyperopia, but further optimisation and validation are needed before its role in screening programmes can be established.

## Data Availability

All data produced in the present study are available upon reasonable request to the authors

## Acknowledgements

The authors are grateful to all participants for their time and to the school administration for their hospitality. Many thanks to optometry students at USN for their help in collecting data, to Stuart J. Gilson for support with software and data management, and to Kim Robert Gustavsen for assistance with scanning electron microscopy measurements.

